# Clinical evaluation of an automated, rapid mariPOC^®^ antigen test in screening of symptomatics and asymptomatics for SARS-CoV-2 infections

**DOI:** 10.1101/2022.04.22.22273686

**Authors:** Marianne Gunell, Kaisa Rantasärkkä, Rita Arjonen, Antti Sandén, Tytti Vuorinen

**Affiliations:** Clinical Microbiology, Laboratory Division, Turku University Hospital; Institute of Biomedicine, University of Turku; Kaarina City Health Care Center

## Abstract

A novel automated mariPOC® SARS-CoV-2 antigen test was evaluated in a Health care center laboratory among symptomatic and asymptomatic individuals seeking SARS-CoV-2 testing. According to the national testing strategy, RT-PCR was used as a reference method. A total of 962 subjects were included in this study, 4.8% (46/962) of their samples were SARS-CoV-2 RT-PCR positive, and 87% (40/46) of these were from symptomatics. Among the symptomatics, the overall sensitivity of the mariPOC® SARS-CoV-2 test was 82.5% (33/40), though the sensitivity increased to 97.1% (33/34) in samples with a Ct value <30. The mariPOC® SARS-CoV-2 test detected 2/6 PCR positive samples among the asymptomatics, four cases that remained antigen test negative had Ct values between 28 and 36. The specificity of the mariPOC® SARS-CoV-2 test was 100% (916/916). The evaluation showed that the mariPOC® SARS-CoV-2 rapid antigen test is very sensitive and specific for the detection of individuals who most probably are contagious.

## Introduction

The global COVID-19 pandemic caused by severe acute respiratory syndrome coronavirus 2 (SARS-CoV-2) has been a significant burden for both society and the carrying capacity of health care since late 2019 [1] when this emerging virus was initially recognized in Wuhan, China. SARS-CoV-2 can mutate into the new emerging variants escaping immunity, and it can in addition to symptomatic infections, manifest as both asymptomatic and presymptomatic infections, and thus the virus has spread efficiently all over the world. According to a simulation model made by US CDC, transmission from asymptomatic individuals, including presymptomatic individuals and those who never develop symptoms, is estimated to account for more than half of all SARS-CoV-2 infections [2]. To prevent the spread of infection there is a need for rapid and accurate diagnostic tests, which detect contagious individuals irrespective of their presence or absence of COVID-19 symptoms.

PCR-based methods, especially RT-PCR is up to date considered the cornerstone for fighting against the pandemic [3,4]. However, large-scale RT-PCR testing, although with excellent sensitivity and analytical specificity, also has some major disadvantages such as long turnaround time as well as the requirement for sophisticated equipment and highly trained personnel. Furthermore, it has been proposed that a positive PCR result may not correlate with infectivity [5], as viral nucleic acids can be detected for a long time after the acute infection, without the presence of infectious and actively replicating SARS-CoV-2 virus [6-12]. As the COVID-19 continues to be a worldwide threat, there is a continuous demand for rapid testing of SARS-CoV-2. Several inexpensive and easy-to-use rapid antigen tests have been developed [13]. Rapid antigen tests have been shown to correlate more accurately with SARS-CoV-2 viral culture than RT-PCR [12], thus also controversial results have been reported [5,14]. Rapid antigen testing of SARS-CoV-2 as a complementary diagnostic method alongside RT-PCR testing has been recently accepted [3,4,15]. ECDC has recommended the use of antigen tests for SARS-CoV-2 diagnostics with a sensitivity of at least 80% and specificity of at least 97% [16].

The technique of the mariPOC^®^ SARS-CoV-2 rapid antigen test (ArcDia International Ltd, Finland) is based on the detection of conserved SARS-CoV-2 nucleocapsid protein with specific monoclonal antibodies. Most positive test results are reported after 20 minutes and final results within 55 minutes [17,18]. The mariPOC^®^ platform is an automated and random access test system that enables simple and quick workflow, high capacity testing, as well as objective result readout. On the platform, the SARS-CoV-2 test is also available as part of syndromic multianalyte tests Quick Flu+ (20 minutes results only) and Respi+ (final results in two hours).

In the present study, the clinical performance of the mariPOC^®^ SARS-CoV-2 rapid antigen test was prospectively evaluated in samples collected in the city of Kaarina, Southwest Finland during spring 2021. Results of on-site testing were compared with central laboratory RT-PCR results to estimate the clinical sensitivity and specificity of the mariPOC^®^ SARS-CoV-2 antigen test.

## Materials and methods

### Study population and specimen collection

An automated mariPOC^®^ SARS-CoV-2 antigen test system was verified for use in the Kaarina city Health care center laboratory for SARS-CoV-2 diagnostics. Verification was conducted between February and May 2021, when the prevalence of SARS-CoV-2 positivity among the tested samples in South-West Finland was approximately 4%. The main circulating SARS-CoV-2 variant in the geographical area during the study period was the Alpha variant (B.1.1.7). At the time, according to the Finnish national COVID-19 hybrid strategy, all individuals having respiratory symptoms as well as those exposed to SARS-CoV-2 were tested and screened, respectively, for SARS-CoV-2.

Two consecutive nasopharyngeal swab (NPS) specimens were obtained from a total of 939 subjects after collecting oral consent. Age, gender, symptoms, and time from the symptom onset were collected from each subject. Of the subjects, 881 had COVID-19-like symptoms and 58 were asymptomatic. The first collected NPS specimen was placed into a viral transport medium (VTM, Bioer sample preservative fluid, BSC82X1-A1) and transported to Turku University Hospital for SARS-CoV-2 RT-PCR testing (cohort 1). The Clinical microbiology laboratory at Turku University Hospital is the primary laboratory responsible for SARS-CoV-2 testing in the Hospital District of Southwest Finland. The second NPS specimen was stored, if needed before mariPOC^®^ analysis, at +4°C in the health care center laboratory.

During the study period, the prevalence of SARS-CoV-2 in the target population was very low. It became soon obvious that based on national verification guidelines of microbiological CE marketed tests [19], a sufficient amount of positive samples to assess test accuracy before introduction in clinical diagnostics, could not be collected in a reasonable time. Therefore, the protocol for sample collection and analysis was altered. Thereafter, together with the strategy implemented in cohort 1, the primary screening of SARS-CoV-2 positive samples was performed with RT-PCR in the Clinical Microbiology laboratory. PCR positive samples were stored at -20 °C and later analyzed by mariPOC^®^ antigen test in the Health care center laboratory (cohort 2). Of the samples in cohort 2, two were omitted from the analysis due to improper handling of the samples before being aliquotted for mariPOC^®^ testing and thus 23 consecutive SARS-CoV-2 positive samples of which six were taken from asymptomatic subjects, were included. For this cohort, NPS specimens were suspended into 2 ml VTM (VACUETTE^®^ Virus Stabilization tube, 456162) for the primary screening of SARS-CoV-2 by RT-PCR. In contrast to Bioer tube, VACUETTE VTM was found to be applicable also in mariPOC^®^ antigen analysis.

### *In-house* SARS-CoV-2 RT-PCR

SARS-CoV-2 RT-PCR from NPS specimens was performed in the Clinical microbiology laboratory at Turku University Hospital. Nucleic acid extraction was performed with Chemagic™ 360 extractor with Viral DNA/RNA 300 Kit H96 (PerkinElmer, Turku, Finland). The in-house RT-PCR test used for SARS-CoV-2 E gene detection was based on the Charité protocol by Corman et al. [20]. The human β-actin gene was used as an internal control (IC) in the test. Final primer concentrations were 400 nM for E gene primers and 200 nM for E gene probe, 40nM for β-actin primers bA-F926 5’-TTGCCGACAGGATGCAGAA-3’ and bA-P954 5’-TGCCCTGGCACCCAGCACAA-3’ and 80 nM for probe bA-R1001 5’-HEX-TCAGGAGGAGCAATGATCTTGAT-BHQ-1-3’. SensiFAST™ Probe No-ROX One-Step Kit (Meridian Bioscience, USA) was used for RT-PCR. Each 25 μL reaction consisted of 12.5 μL of 2X SensiFAST Probe One-Step mix, 1 μL of E gene primers and 0.5 μL of E gene probe, 0.1 μL of β-actin primers, and 0.2 μL β-actin probe, 0.2 μL Reverse transcriptase, 0.4 μL RiboSafe RNase Inhibitor and 9 μL of extracted RNA template. Cycling conditions were 55 °C (10 min), 95 °C (3 min) followed by 45 cycles of 95 °C (15 s) and 58 °C (30 s) performed with BMS MIC analyzers (BMS Australia).

### mariPOC^®^ SARS-CoV-2 antigen test

The mariPOC^®^ SARS-CoV-2 testing was performed in the on-site laboratory of Kaarina Health Care Center. NPS specimens from cohort 1 were suspended into 1.3 ml mariPOC^®^ RTI sample buffer in sample tubes and analyzed with the mariPOC^®^ test system according to the manufacturer’s instructions as soon as possible. The samples in cohort 2 were collected in VACUETTE VTM and stored at -20 °C after the primary SARS-CoV-2 PCR test and were further diluted 1:1 (0.65 ml+ 0.65 ml) with mariPOC^®^ RTI sample buffer to gain the required sample volume for mariPOC^®^ analysis. The VTM samples were diluted approximately 3-times more than in the dry swab procedure recommended by the mariPOC^®^ manufacturer.

### Statistical analysis

The mariPOC^®^ SARS-CoV-2 rapid antigen test sensitivity, including 95% confidence intervals (CI), was determined using MedCalc Software [21].

## Results

### Study population

Demographic data of the population included in the mariPOC^®^ rapid antigen test evaluation is presented in Table 1. Of the whole study population, 6.7% (64) were asymptomatic and 93.3% (898) had symptoms linked to COVID-19, such as sore throat, headache, fever, shortness of breath, and diarrhea.

**Table 1.** Demographic data on the population included in the mariPOC^®^ SARS-CoV-2 rapid antigen test evaluation study

| Demographic data | Female | Male |
| --- | --- | --- |
| Sex distribution (%) | 575 (59.8) | 387 (40.2) |
| Median age | 40.9 years | 39.2 years |
| Age distribution | 4-81 years | 2-81 years |
| <18 year of age (%) | 9 (0.9) | 20 (2.1) |
| >65 years of age (%) | 30 (3.1) | 20 (2.1) |
| SARS-CoV-2 positive (%) | 22 (47.8) | 24 (52.2) |

### SARS-CoV-2 RT-PCR test results

Totally, 962 samples were analyzed with RT-PCR. Of the tested samples, 46 (4.8%) were SARS-CoV-2 positive with the RT-PCR method. Ct values for E gene amplification varied from 14.66 (high RNA load) to 38.13 (low RNA load). Ct values <40 cycles for the E gene were interpreted as SARS-CoV-2 positive. In cohort 1, all 23 subjects with RT-PCR positivity had COVID-19 symptoms, whereas six of the 23 PCR positive samples in cohort 2 were from asymptomatic subjects and 17 subjects had COVID-19-like symptoms.

### Comparison of mariPOC^®^ SARS-CoV-2 antigen test and RT-PCR test results

The correlation of Ct values and mariPOC^®^ SARS-CoV-2 rapid antigen test results among asymptomatic and symptomatic subjects are presented in Figure 1. Totally, 35 out of 46 (76.1%) of the SARS-CoV-2 RT-PCR positive samples were positive in the mariPOC^®^ test (true positive). The overall sensitivity for symptomatic patients including both cohorts 1 and 2 was 97.1% (33/34) and 82.5% (33/40) when Ct values <30 and <40 were used, respectively (Table 2). The mariPOC^®^ test was positive for up to 10 days from the onset of symptoms.

**Table 2.** Clinical sensitivity and 95% confidence intervals of mariPOC^®^ SARS-CoV-2 rapid antigen test among symptomatic subjects at 20 minutes (preliminary) and 55 minutes (final) outcome in correlation to Ct values of the reference RT-PCR method.

|  |  | mariPOC® positives |  | PCR positives | mariPOC® Sensitivity (95% CI) |  |
| --- | --- | --- | --- | --- | --- | --- |
|  |  | 20 min outcome | 55 min outcome |  | 20 minutes outcome | 55 minutes outcome |
| **Cohort 1** | Ct <30 | 17 | 18 | 18 | 94.4% (72.7-99.9%) | 100.0% (81.5-100%) |
|  | Ct <33 | 17 | 18 | 20 | 85.0% (62.1-96.8%) | 90.0% (68.3-98.8%) |
|  | Ct <40 | 17 | 18 | 23 | 73.9% (51.6-89.8%) | 78.3% (56.3-92.5%) |
| **Cohort 2** | Ct <30 | 13 | 15 | 16 | 81.3% (54.4-96.0%) | 93.8% (69.8-99.8%) |
|  | Ct <33 | 13 | 15 | 17 | 76.5% (50.1-93.2%) | 88.2% (63.6-98.5%) |
|  | Ct <40 | 13 | 15 | 17 | 76.5% (50.1-93.2%) | 88.2% (63.6-98.5%) |
| **Cohort 1 & 2** | Ct <30 | 30 | 33 | 34 | 88.2% (72.6-96.7%) | 97.1% (84.7-99.9%) |
|  | Ct <33 | 30 | 33 | 37 | 81.1% (64.8-92.0%) | 89.2% (74.6-97.0%) |
|  | Ct <40 | 30 | 33 | 40 | 75.0% (58.8-87.3%) | 82.5% (67.2-92.7%) |

**Figure 1.**
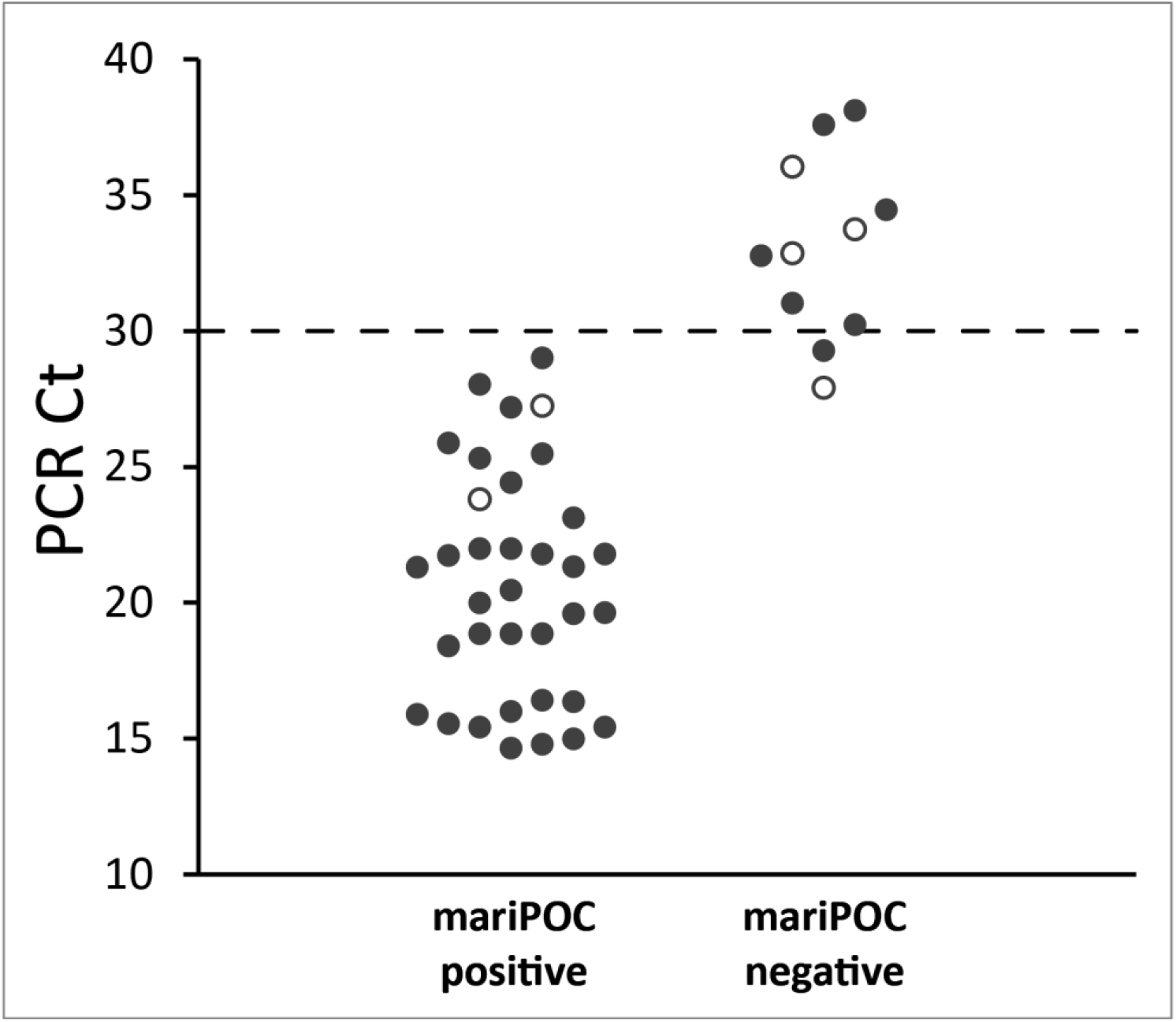
Correlation of Ct values of RT-PCR method and mariPOC^®^ SARS-CoV-2 rapid antigen test among symptomatic (black dots) and asymptomatic (empty dots) subjects.

In cohort 1, 18 PCR-positive samples were positive in the mariPOC^®^ SARS-CoV-2 rapid antigen test and five samples remained negative. Ct values for 18 true positive samples in the antigen detection varied from 14.80 to 29.01 (Figure 1) and the mean duration of symptoms was 2.5 days (range 1-10 days). Of the five false-negative samples in the antigen detection, Ct values varied from 30.24 to 38.13, and the mean duration of symptoms was two days (range 0-5 days). The mariPOC^®^ test sensitivity for cohort 1 was 78.3% (18/23, Table 2). When only samples with Ct values <30 were considered, the mariPOC^®^ sensitivity in cohort 1 within symptomatic subjects was 100.0% (18/18).

In cohort 2, 17 of the 23 PCR positive samples were mariPOC^®^ SARS-CoV-2 test positive and six samples remained negative (Table 2). Four of the samples from cohort 2 were taken from patients whose second NPS specimen was also included in cohort 1. Of the six PCR positive samples that remained negative in the antigen detection, four were taken from asymptomatic patients and two have had only mild COVID-19 symptoms for one or two days. Ct values for 17 true positive samples in the antigen detection varied from 14.66 to 27.25 (Figure 1) and the mean duration of symptoms was 2.5 days (range 0-7 days). Ct values for the false-negative samples in the antigen detection varied from 27.91 to 36.06, only two samples had Ct value <30 (Figure 1), and the mean duration of symptoms was 0.5 days (range 0-2 days). The sensitivity of the mariPOC^®^ antigen test for cohort 2 was 73.9% (17/23), but when only symptomatic subjects were considered, the sensitivity was 88.2% (15/17, Table 2).

The results of the mariPOC^®^ SARS-CoV-2 test reported after 20 minutes correlated well with the final results. Only three samples (one in cohort 1 and two in cohort 2) were negative after 20 minutes and turned into positive in final results at 55 minutes outcome. The Ct values for these positive samples varied from 24.43 to 28.04, and the duration of symptoms was 1–5 days.

## Discussion

Early and accurate detection of SARS-CoV-2 infection is crucial for reducing virus transmission in the community. During the COVID-19 pandemic, the need for rapid testing has raised significantly and a high number of antigen tests have been introduced on the market. mariPOC^®^ is a fully automated test system that enables the testing of up to 100 samples within a work shift at the sampling site. Over 90% of SARS-CoV-2 positive results are obtained in 20 minutes and low positive and negative results are reported after 55 minutes. Hands-on time is short and analysis, as well as result reading, is automated. These properties make mariPOC^®^ test systems suitable for use in medium and small-size volume laboratories and decentralized testing.

The evaluation of the mariPOC^®^ SARS-CoV-2 rapid antigen test to be used in a health care laboratory was performed in a medium-sized Finnish city representing adequate variation in social, ethnic, and age distribution of the population seeking COVID-19 testing in Finland. The SARS-CoV-2 positivity rate among the study population was 4.8%, determined by the RT-PCR method, which was well in correlation to the SARS-CoV-2 prevalence in the hospital district of Southwest Finland during spring 2021.

The sensitivities of antigen tests in different published studies have varied considerably due to differences in evaluated test products, study protocols, and patient cohorts [22,23]. The overall sensitivity of the mariPOC^®^ antigen test in our evaluation was 76.1% which is in correlation with a recent meta-analysis showing the overall pooled sensitivity of 72.1% of the antigen tests in publications fulfilling the criteria to be included in the meta-analysis [24]. When the results of cohorts 1 and 2 were assessed separately, the sensitivity in cohort 2 was lower (73.9%) than that in cohort 1 (78.3%). This could be explained by the fact that in cohort 2, four out of six false-negative samples were from asymptomatic subjects who most probably carry less SARS-CoV-2 virus than the symptomatic subjects. Threshold cycle (Ct) data from our RT-PCR was in line with this proposition. Furthermore, in cohort 2, the samples in VACUETTE tubes were diluted 3-times more for mariPOC^®^ analysis compared to the recommended procedure of the manufacturer (NPS collected directly in mariPOC^®^ RTI sample buffer). No false-positive findings were reported in this study.

The performance of antigen tests to detect SARS-CoV-2 is known to be highest during the first seven days from the appearance of symptoms [23,34] and most guidelines advise using rapid tests accordingly and in symptomatic subjects [4,16]. Our results show, that the overall sensitivity of 82.5% was reached when the sensitivity for samples obtained only from symptomatic individuals was assessed, thus showing the good performance of the mariPOC^®^ SARS-CoV-2 test in diagnostic testing. While the number of positive samples among asymptomatic individuals was low (n=6), definitive conclusions about the use of the mariPOC^®^ antigen test in detecting SARS-CoV-2 in asymptomatics cannot be drawn based on this study.

Infectivity of SARS-CoV-2 is associated with viral load and the lower Ct values in RT-PCR indicate higher viral load [25]. In the present study, we have shown that asymptomatic RT-PCR positive subjects had on average higher Ct values compared to subjects with symptoms and that antigen test sensitivity increases when Ct values decrease. When Ct <30 was used as a threshold, the sensitivity of the antigen test was 97.1% and even up to 100% when only symptomatic patients were included. Thus, the sensitivity of the mariPOC^®^ rapid antigen test correlates better to RT-PCR Ct value than the patient symptom status or the intensity of symptoms, indicating that the mariPOC^®^ SARS-CoV-2 rapid antigen test recognizes well the subjects with contagious SARS-CoV-2 infection [26-28].

Results for the SARS-CoV-2 test of the mariPOC^®^ test system are reported in two phases. At 20 minute outcome, most positive results are reported and due to the high specificity (here 100%), those results are reliable. At 55 minutes outcome also low positives and negatives are reported. In our study, 91.4% (32/35 total mariPOC^®^ antigen test positives) of positive samples were positive already at 20 minutes. Our prospective evaluation results of the mariPOC^®^ test are well in line with those reported earlier from a retrospective study [18].

Although antigen tests have lower sensitivity compared to RT-PCR methods [25,29], to fight against COVID-19 pandemics both PCR and antigen tests are needed [5,14,15]. Especially in places where central hospital laboratory facilities are not available, shorter turnaround time and ease of use make antigen tests a powerful tool to prevent the spread of COVID-19. In addition, rapid antigen tests, such as automated mariPOC^®^ SARS-CoV-2, could be a good alternative for large-scale screening of individuals at schools and workplaces and, therefore, help to prevent the spread of the COVID-19 in the community [28,30].

## Conclusions

We conclude that the mariPOC^®^ SARS-CoV-2 antigen test detected the majority of the samples with RT-PCR cycle threshold below 30 among symptomatic and asymptomatic subjects justifying its use for rapid detection of individuals who most probably are contagious. In addition, the mariPOC^®^ test system is practicable in small and medium-size laboratories as well as health care centers to be used for rapid SARS-CoV-2 detection in symptomatics.

## Data Availability

All data produced in the present work are contained in the manuscript

## Transparency declaration

### Research permission and ethics declaration

This study was approved by the Hospital District of Southwest Finland, research approval number T12/009/21. Ethical aspects of the study were considered and approved by Turku Clinical Research Centre (Turku CRC), no ethical committee review was required. Oral consent for sample collection was obtained from each person. The samples were taken to apply a new test method for SARS-CoV-2 diagnosis in the laboratory and the results between the two tests were anonymously processed

## Acknowledgements

Heta-Maija Manner and her colleagues (Kaarina city Health care center) are thanked for their skillful technical assistance with mariPOC^®^ analyses.

